# Confounding adjustment performance of ordinal analysis methods in stroke studies

**DOI:** 10.1101/19000943

**Authors:** T.P. Zonneveld, A. Aigner, R.H.H. Groenwold, A. Algra, P.J. Nederkoorn, U. Grittner, N.D. Kruyt, B. Siegerink

## Abstract

**Background:** In acute stroke studies, ordinal logistic regression (OLR) is often used to analyze outcome on the modified Rankin Scale (mRS), whereas the non-parametric Mann-Whitney measure of superiority (MWS) has also been suggested. It is unclear how these perform comparatively when confounding adjustment is warranted. Our aim is to quantify the performance of OLR and MWS in different confounding variable settings.

**Methods:** We set up a simulation study with three different scenarios; (1) dichotomous confounding variables, (2) continuous confounding variables, and (3) confounding variable settings mimicking a study on functional outcome after stroke. We compared adjusted ordinal logistic regression (aOLR) and stratified Mann-Whitney measure of superiority (sMWS), and also used propensity scores to stratify the MWS (psMWS). For comparability, OLR estimates were transformed to a MWS. We report bias, the percentage of runs that produced a point estimate deviating by more than 0.05 points (point estimate variation), and the coverage probability.

**Results:** In scenario 1, there was no bias in both sMWS and aOLR, with similar point estimate variation and coverage probabilities. In scenario 2, sMWS resulted in more bias (0.04 versus 0.00), and higher point estimate variation (41.6% versus 3.3%), whereas coverage probabilities were similar. In scenario 3, there was no bias in both methods, point estimate variation was higher in the sMWS (6.7%) versus aOLR (1.1%), and coverage probabilities were 0.98 (sMWS) versus 0.95 (aOLR). With psMWS, bias remained 0.00, with less point estimate variation (1.5%) and a coverage probability of 0.95.

**Conclusions:** The bias of both adjustment methods was similar in our stroke simulation scenario, and the higher point estimate variation in the MWS improved with propensity score based stratification. The stratified MWS is a valid alternative for adjusted OLR only when the ratio of number of strata versus number of observations is relatively low, but propensity score based stratification extends the application range of the MWS.

## Introduction

The ordinal *modified Rankin Scale* (mRS) measures functional outcome after stroke on a 7-step scale from 0 (no symptoms) to 6 (death), and is the primary outcome measure in most acute stroke trials.(1) To analyze the differences in mRS between treatment arms, pivotal stroke trials primarily use the ordinal logistic regression (OLR) method.(2-4) The OLR produces a single effect size estimate (*a common odds ratio*) based on the odds ratios for each cut-point across the mRS, and this estimate can be interpreted as the odds ratio of ending up in a higher category of the scale. But as OLR is based on several assumptions, such as a linear and proportional effect of the independent variables on the outcome variable,(5) the Mann-Whitney measure of superiority (MWS) was recently proposed as a more robust analysis method of an ordinal outcome scale.(6) In contrast to regression methods, the MWS is a non-parametric rank-based test based on *proversions*, which are one to one comparisons of outcome between observations. In short, each observation in one group (A) is compared to each observation in the other group (B), and the following three complementary probabilities (Ps) are derived: P(A>B), P(A=B), and P(B>A). The MWS for A is then given by the formula P(A>B) + 0.5P(A=B). As a result, the MWS ranges from 0 to 1, with 0.5 as the value of no difference between groups A and B.

Importantly, OLR and MWS differ fundamentally in their confounding adjustment technique. In regression methods such as the OLR, independent variables can be added to the equation. However, these models are also subject to aforementioned assumptions. As a non-parametric method, the MWS uses stratification for confounding adjustment: the basic concept is that the proversions are performed only *within* the defined strata. For example, when adjusting for sex, proversions are only made between males from group A versus males from group B, and females from group A versus females from group B. Stratification is however linked to estimation problems. Most notably, residual confounding and instability through empty cells might occur, especially when adjusting for multiple confounding variables. A possible solution to overcome these issues is to form strata based on percentiles of propensity scores, which estimate the probability of being exposed based on measured confounding variables.(7)

It has previously not been investigated how the different adjustment techniques of OLR and MWS perform comparatively. Our objective was to compare and quantify the bias/variance trade-off of these methods in a simulation model with varying confounding conditions, focusing on those conditions typically present in an acute ischemic stroke cohort.

## Methods

### Scenarios

We generated data in three distinct scenarios, differing from each other only in their confounding variable settings (table 1). In scenario 1, we modeled five *dichotomous* confounding variables all with a prevalence of 0.5 and all with a regression coefficient of ln(1.5) (which is equivalent to an odds ratio of 1.5) in relation to the outcome. In scenario 2, we modeled five *continuous* confounding variables, with a standard normal distribution and regression coefficient of ln(1.5) in relation to the outcome. In scenario 3, we modeled five *varying* confounding variables, with distributions and regression coefficients reflecting known important characteristics (sex, age, stroke severity, previous stroke, systolic blood pressure) associated with functional outcome after stroke.(8, 9) In our main simulations we generated 1000 observations, which we changed to 250 and 4000 in sensitivity analyses. Each scenario was run 1000 times. All simulations were performed in Stata/IC 15.1 for Windows (32 bit), with full code provided in the appendix.

**Table 1.** Confounding variable settings per scenario. Abbreviations: RC = regression coefficient; NIHSS = National Institutes of Health Stroke Scale; SBP = systolic blood pressure; SD = standard deviation; IQR = interquartile range; ln = natural logarithm. * = per unit increase.

| Scenario | Confounding variables | Type | Distribution | Prevalence | RCs ($\beta_j$) |
| --- | --- | --- | --- | --- | --- |
| 1 | $x_1, x_2, x_3, x_4, x_5$ | Dichotomous | Binary | 50% | $\ln(1.50)$ |
| 2 | $x_1, x_2, x_3, x_4, x_5$ | Continuous | Normal | Mean=0 (SD=1) | $\ln(1.50)$ |
| 3 | $x_1$; female sex | Dichotomous | Binary | 50% | $\ln(1.10)$ |
| | $x_2$; age | Continuous | Normal | Mean=69 (SD=10) | $\ln(1.01)^*$ |
| | $x_3$; NIHSS | Ordinal | Right-skewed | Median=9 [IQR=5-14] | $\ln(1.05)^*$ |
| | $x_4$; previous stroke | Dichotomous | Binary | 50% | $\ln(1.10)$ |
| | $x_5$; SBP | Continuous | Normal | Mean=160 (SD=15) | $\ln(1.01)^*$ |

### Data generation process

First, we generated a seven-step ordinal outcome variable, based on the presence or value of the confounding variables and their assumed relationship with the outcome (as specified in the respective scenario). Second, we constructed a dichotomous exposure variable also based on the confounding variables present, yet conditionally independent of the outcome. Importantly, we did not model a direct relationship between the exposure variable and the ordinal outcome variable, nor did we model a correlation between any of the confounding variables. See appendix 1 for a detailed description of our data generation process and appendix 2 for the full Stata code used.

### Comparison of analysis methods

For each run, we performed a crude OLR and MWS analysis, and an adjusted analysis for both methods; regression adjustment in OLR (aOLR) and stratified adjustment in MWS (sMWS). Ordinal and continuous confounding variables were stratified based on quartiles; this resulted in up to 32 (2^5) possible strata in scenario 1, up to 1024 (4^5) possible strata in scenario 2, and up to 256 (2*4*4*2*4) possible strata in scenario 3. We calculated a propensity score per observation based on all confounding variables present in the respective scenario. This score was subsequently divided into quartiles for the propensity based stratified MWS (psMWS). For comparison purposes, we converted the odds ratios (ORs) generated by the OLR to Mann-Whitney measures of superiority, with the following approximation formula: *MWS = (OR/(OR-1)*^*2*^*) × ((OR-1)-ln(OR))*.(6)

### Outcome parameters

The validity of each method is assessed by the extent of the bias, which we defined as the difference between the mean of the observed point estimates and the simulated, true effect. In order to quantify the variation in (point) estimates of each method, we report the percentage of runs that produced a point estimate deviating more than 0.05 MWS from the true effect (i.e. an estimate lower than 0.45 or higher than 0.55, roughly equivalent to an OR lower than 0.74 or higher than 1.35 using the approximation formula stated above). Finally, we also report the coverage probability, defined as the proportion of 95%-confidence intervals encompassing the true effect. With our 1000 runs, calculated coverage probabilities within the range of 93.6% - 96.4% are compatible with a true coverage of 95%. Of note, as the number of *proversions* decreases when the number of strata increases, there were no or only very little *proversions* to construct the MWS estimate in some runs. As this results in extreme estimates and impossibility to construct a valid confidence interval, we discarded runs that resulted in less than 11 *proversions*. We created boxplots of the five analysis methods’ (OLR, MWS, aOLR, sMWS, psMWS) point estimates, displaying the lower adjacent value, 25^th^ percentile, median, 75^th^ percentile, and upper adjacent value (extreme outliers not shown).

## Results

### Scenario 1: Dichotomous confounding variables

In the scenario with five dichotomous confounding variables (resulting in 32 possible strata for sMWS), sMWS and aOLR performed similar; bias was 0.00 in both methods, and point estimate variation (percentage of runs that produced a point estimate deviating more than 0.05 MWS from the true effect) was 2.1% in the sMWS versus 1.8% in the aOLR. The coverage probability was 96% in the sMWS versus 95% in the aOLR. Propensity score based strata adjustment in the MWS (psMWS) resulted in a bias of 0.01, a point estimate variation of 2.1%, and coverage probability of 93%. See figure 1 for the boxplots (including the results for 1 to 4 dichotomous confounding variables).

**Figure 1.**
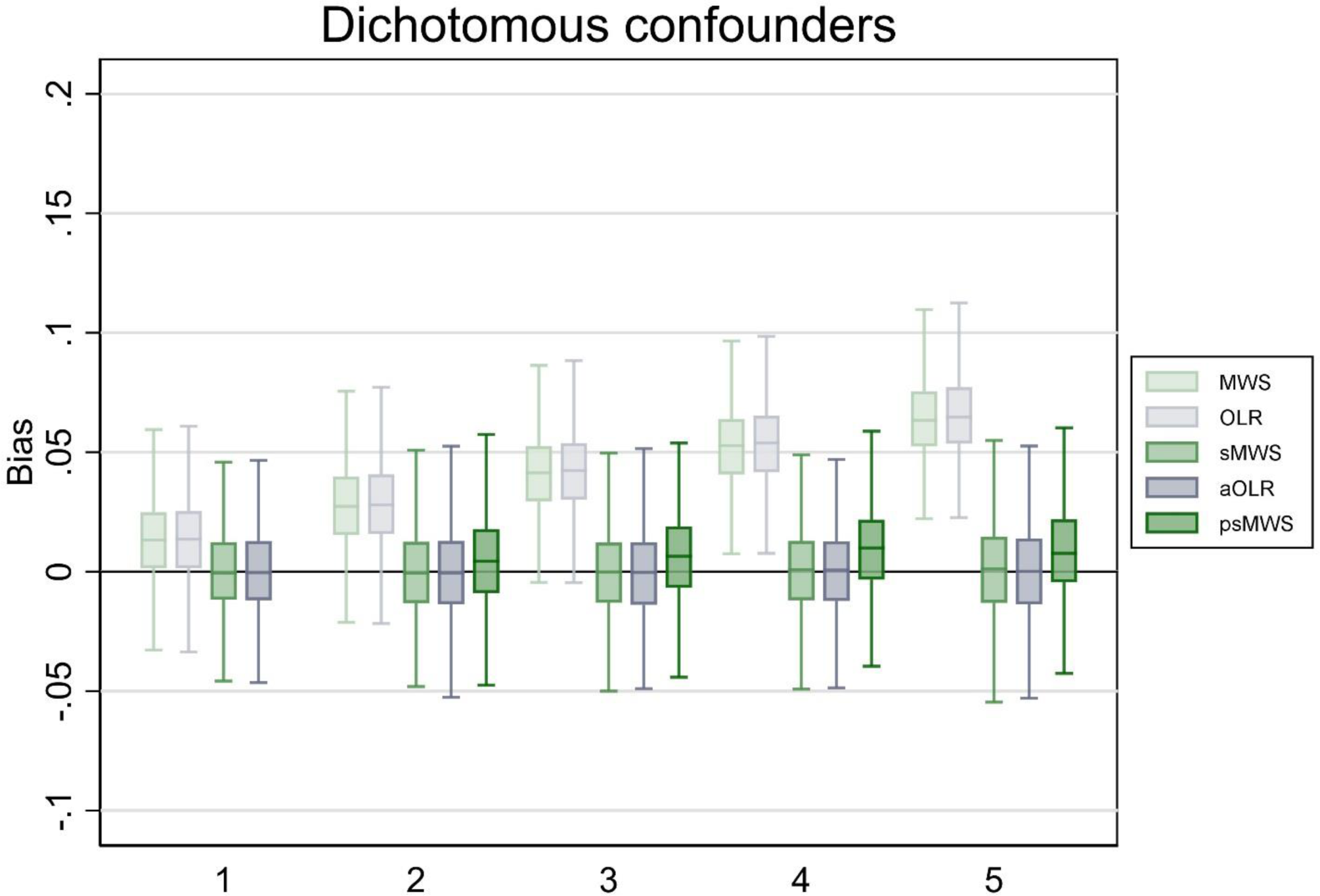
Bias of Mann-Whitney measure of superiority (MWS), ordinal logistic regression (OLR), stratified Mann-Whitney measure of superiority (sMWS), adjusted ordinal logistic regression (aOLR), and propensity score based stratified Mann-Whitney measure of superiority (psMWS) in scenario 1. The psMWS was not performed in the scenario with one confounding variable. Runs (N): 1000. The x-axis shows the number of confounding variables modeled. The y-axis shows the bias, with estimates from the OLR analyses converted to a MWS. Boxplots display the lower adjacent value, 25^th^ percentile, median, 75^th^ percentile, and upper adjacent value (extreme outliers are not displayed).

### Scenario 2: Continuous confounding variables

In the scenario with five continuous confounding variables (resulting in 1024 possible strata for sMWS), aOLR outperformed sMWS; bias was 0.04 in the sMWS versus 0.00 in the aOLR, and point estimate variation was 41.6% in the sMWS versus 3.3% in the aOLR. The coverage probability was 96% for both methods. With psMWS, bias was 0.02, point estimate variation was 8.1%, and coverage probability was 88%. See figure 2 for the boxplots (including the results for 1 to 4 continuous confounding variables).

**Figure 2.**
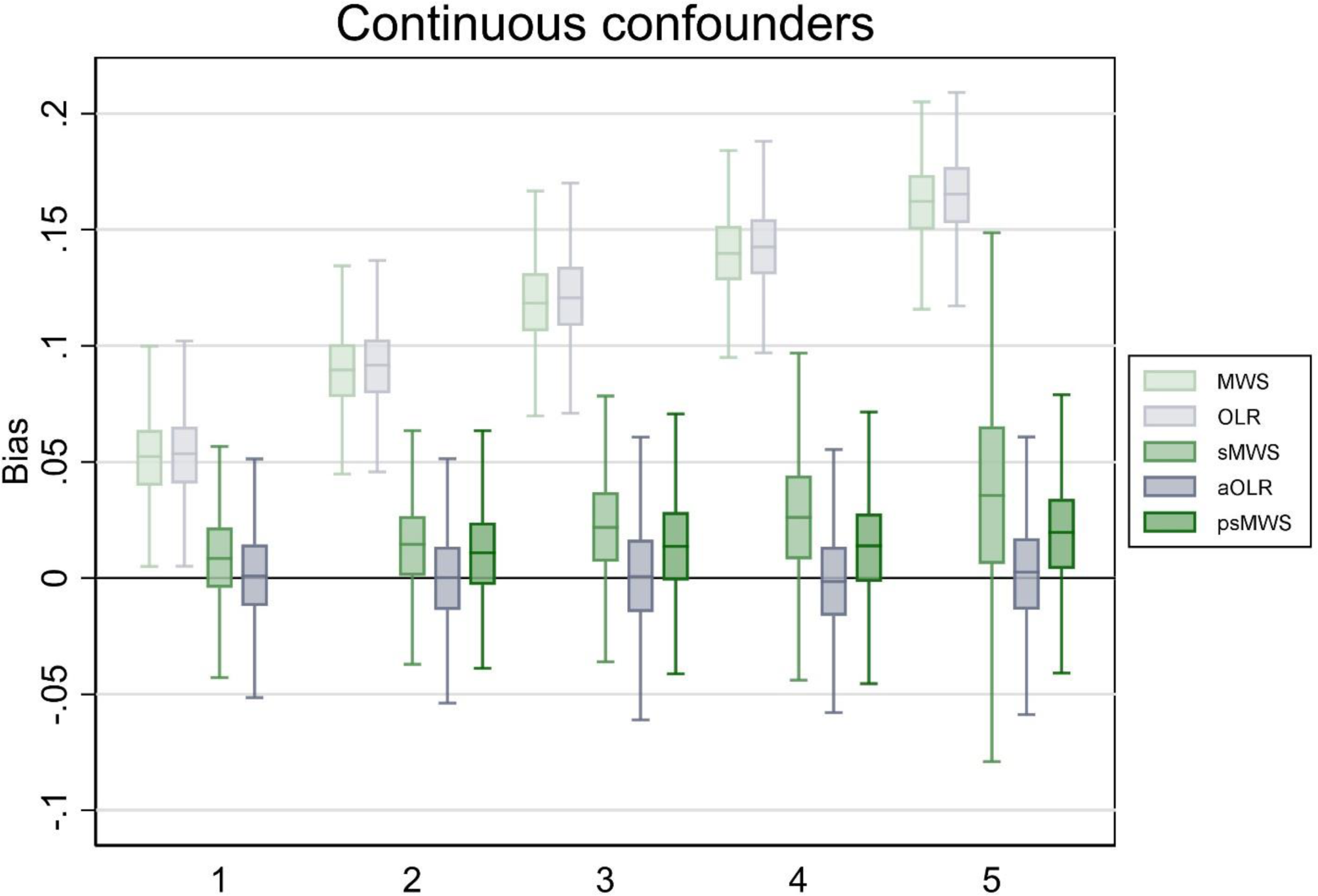
Bias of Mann-Whitney measure of superiority (MWS), ordinal logistic regression (OLR), stratified Mann-Whitney measure of superiority (sMWS), adjusted ordinal logistic regression (aOLR), and propensity score based stratified Mann-Whitney measure of superiority (psMWS) in scenario 2. The psMWS was not performed in the scenario with one confounding variable. Runs (N): 1000. The x-axis shows the number of confounding variables modeled. The y-axis shows the bias, with estimates from the OLR analyses converted to a MWS. Boxplots display the lower adjacent value, 25^th^ percentile, median, 75^th^ percentile, and upper adjacent value (extreme outliers are not displayed).

### Scenario 3: Varying confounding variables

In the scenario with five varying confounding variables (resulting in 256 possible strata for sMWS), sMWS and aOLR performed similar; bias was 0.00 in both methods, and point estimate variation was 6.7% in the sMWS versus 1.1% in the aOLR. The coverage probability was 98% for sMWS and 95% in the aOLR. With psMWS, bias was 0.00, point estimate variation was 1.5%, and coverage probability was95%. See figure 3 for the boxplots (including the results for 1 to 4 varying confounding variables).

**Figure 3.**
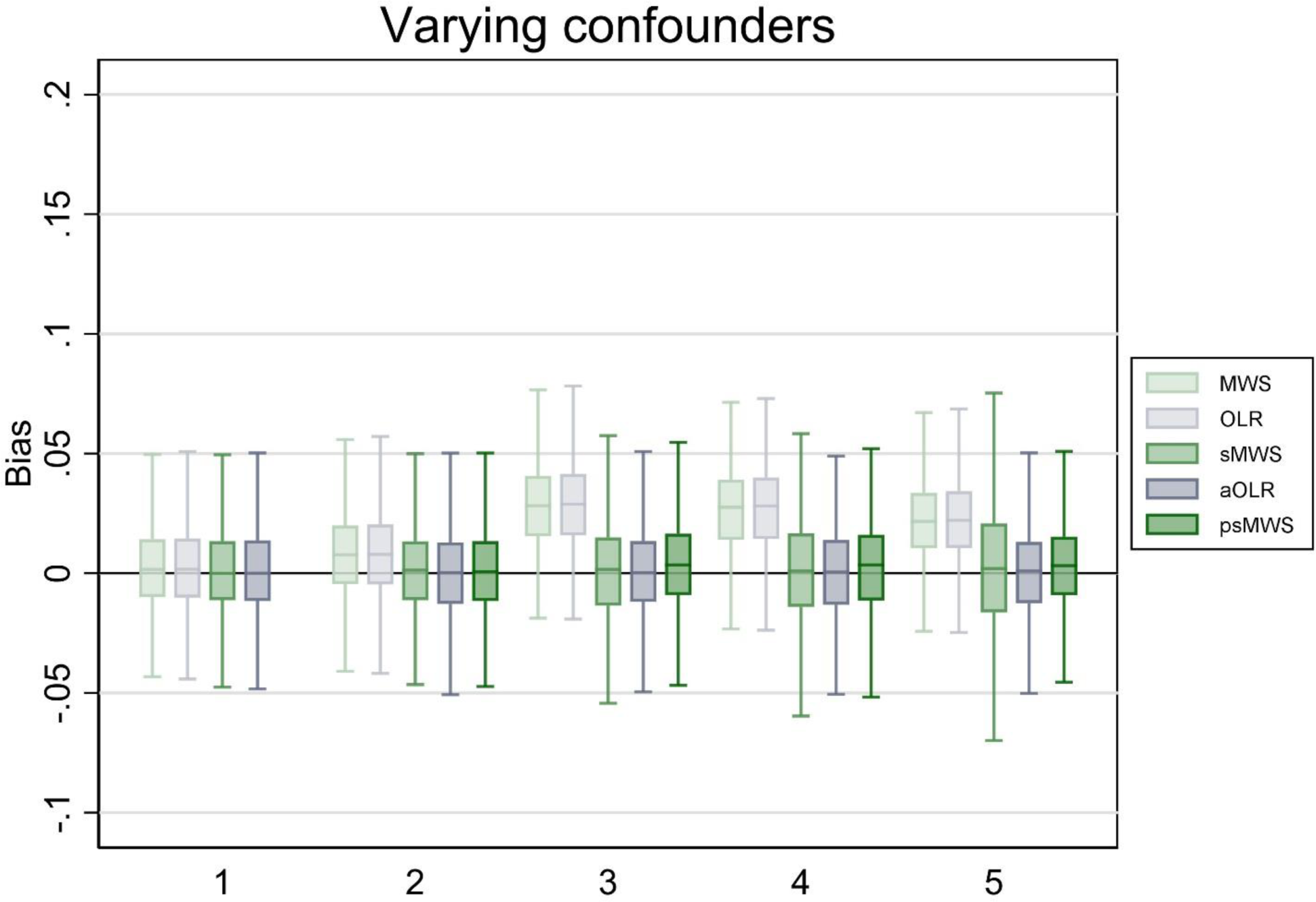
Bias of Mann-Whitney measure of superiority (MWS), ordinal logistic regression (OLR), stratified Mann-Whitney measure of superiority (sMWS), adjusted ordinal logistic regression (aOLR), and propensity score based stratified Mann-Whitney measure of superiority (psMWS) in scenario 3. The psMWS was not performed in the scenario with one confounding variable. Runs (N): 1000. The x-axis shows the number of confounding variables modeled. The y-axis shows the bias, with estimates from the OLR analyses converted to a MWS. Boxplots display the lower adjacent value, 25^th^ percentile, median, 75^th^ percentile, and upper adjacent value (extreme outliers are not displayed).

### Varying sample size (table 2)

Sensitivity analyses with 250 observations in scenario 1 resulted in similar bias (sMWS 0.00, aOLR 0.00, psMWS 0.01) and coverage probabilities (sMWS 96%, aOLR 95%, psMWS 96%) as the main analyses, but with higher point estimate variation (sMWS 30.5%, aOLR 25.0%, psMWS 23.7%). In scenario 2, bias was also similar to the main analyses (sMWS 0.05, aOLR 0.00, psMWS 0.02), but point estimate variation increased particularly in the sMWS (91.0% versus 32.2% with aOLR, and 34.5% with psMWS). Coverage probabilities were 100% (sMWS) versus 95% (aOLR), and 93% in the psMWS. In scenario 3, bias was similar to the main analyses (sMWS 0.01, aOLR 0.00, psMWS 0.00), but point estimate variation increased particularly in the sMWS (58.3% versus 22.5% with aOLR, and 20.4% with psMWS). Coverage probabilities were 98% (sMWS) versus 94% (aOLR), and 96% in the psMWS.

**Table 2.** Varying sample sizes (sensitivity analyses). Results shown for scenarios with five confounding variables. Abbreviations: sMWS = stratified Mann-Whitney measure of superiority; aOLR = adjusted ordinal logistic regression; psMWS = propensity score based stratified Mann-Whitney measure of superiority; PEV = point estimate variation (percentage of runs in which the difference between point estimate and true effect is more than 0.05 MWS); CP = coverage probability.

| Scenario | Outcome | 250 |  |  | 1000 |  |  | 4000 |  |  |
| --- | --- | --- | --- | --- | --- | --- | --- | --- | --- | --- |
|  |  | sMWS | aOLR | psMWS | sMWS | aOLR | psMWS | sMWS | aOLR | psMWS |
| 1 | Bias | 0.00 | 0.00 | 0.01 | 0.00 | 0.00 | 0.01 | 0.00 | 0.00 | 0.01 |
|  | PEV (%) | 30.5 | 25.0 | 23.7 | 2.1 | 1.8 | 2.1 | 0.0 | 0.0 | 0.0 |
|  | CP | 0.96 | 0.95 | 0.96 | 0.96 | 0.95 | 0.93 | 0.95 | 0.95 | 0.86 |
| 2 | Bias | 0.05 | 0.00 | 0.02 | 0.04 | 0.00 | 0.02 | 0.03 | 0.00 | 0.02 |
|  | PEV (%) | 91.0 | 32.2 | 34.5 | 41.6 | 3.3 | 8.1 | 14.7 | 0.0 | 0.1 |
|  | CP | 1.00 | 0.95 | 0.93 | 0.96 | 0.96 | 0.88 | 0.49 | 0.95 | 0.71 |
| 3 | Bias | 0.01 | 0.00 | 0.00 | 0.00 | 0.00 | 0.00 | 0.00 | 0.00 | 0.00 |
|  | PEV (%) | 58.3 | 22.5 | 20.4 | 6.7 | 1.1 | 1.5 | 0.0 | 0.0 | 0.0 |
|  | CP | 0.98 | 0.94 | 0.96 | 0.98 | 0.95 | 0.95 | 0.95 | 0.94 | 0.92 |

Sensitivity analyses with 4000 observations in scenario 1 resulted in similar bias (sMWS 0.00, aOLR 0.00, psMWS 0.01) and coverage probabilities (sMWS 95%, aOLR 95%, psMWS 86%), but, as expected, with lower point estimate variation (all methods 0.0%). In scenario 2, bias was also similar to the main analyses (sMWS 0.03, aOLR 0.00, psMWS 0.02), and point estimate variation lowered proportionally (sMWS 14.7%, aOLR 0.0%, psMWS 0.1%). However, the coverage probability was only 49% with sMWS versus 95% with aOLR, and 71% with psMWS. In scenario 3, bias was 0.00 and point estimate variation was 0.0% in all analysis methods. Coverage probabilities were 95% (sMWS) versus 94% (aOLR), and 92% in the sMWS.

## Discussion

In our simulation scenario with structures of confounding variables based on stroke cohorts (scenario 3), we found that both stratified MWS (sMWS) and adjusted OLR (aOLR) produced unbiased point estimates. The variation in point estimates was higher for sMWS, but this was fixed with propensity score based stratification in the MWS (psMWS). Interestingly, whereas sMWS performed worse than aOLR when modeling fewer observations, but psMWS produced similar results to aOLR. When modeling a larger number of observations, differences disappeared and all methods produced unbiased and precise point estimates. In the scenario with dichotomous confounding variables (scenario 1), both methods performed similar in terms of bias and point estimate variation. In the scenario with continuous confounding variables (scenario 2), sMWS resulted in more bias and higher point estimate variation than aOLR, as can be expected from any stratification based methodology.(10) Although psMWS resulted in improved results compared to sMWS, performance of aOLR remained superior in this scenario.

To our knowledge, this is the first study directly comparing the performance of confounding adjustment of a parametric model (OLR) with a non-parametric test (MWS) regarding bias and precision of resulting effect estimates. Although it is well known that in most cases stratification methods are less effective,(11) we still compared these methods head-to-head as it reflects the choice that stroke researchers have to make in scientific practice. Our comparison was primarily focused on quantifying the differences, to provide researchers with more detailed characteristics of these analyses methods. The MWS seems primarily suited for situations when no adjustment for confounding is indicated, i.e. in primary analyses of interventional studies. But our simulations showed that sMWS also performs comparably to aOLR in a range of confounding variable settings. However, with increasing number of continuous confounding variables (and thus strata), the sMWS becomes more biased and shows higher point estimate variation, which is corrected for only partly by psMWS.

Our simulations have the limitation that they do not address other issues relevant when deciding which analysis technique should be applied. These issues include residual confounding, measurement error and misclassification, model misspecification, and missing data patterns. Although important, we believe these issues are in some sense secondary to the more basic question that we addressed in our simulations. Another limitation is that we modeled proportional effects of our confounding variables on the outcome; further research could focus on exploring the comparative performance of MWS and OLR when violating the *proportional odds* assumption. A final limitation is inseparably linked with the nature of the MWS; as it performs *proversions* between two groups only, it can only be used when studying a binary exposure variable. This might not be a problem in most intervention studies, but dichotomization of a non-dichotomous exposure invariably leads to a loss of information. We firmly believe that the choice on how the exposure is modeled should be based on subject matter knowledge in combination with a weighing of potential drawbacks of analysis techniques. Therefore, we are not able to provide a general statement whether the benefits of the assumption free MWS approach outweigh drawbacks that come from the required categorization of both the exposure and the confounding variables. Other research fields than stroke might have a different constellation of known confounding factors, which renders it difficult to extrapolate our results to other fields. Yet, as we provide the used Stata code, readers could modify the provided code in the appendix to generate results more relevant to their specific setting.

In conclusion, the confounding variable settings in our stroke simulation scenario resulted in an unbiased performance of both methods, and the higher point estimate variation in the stratified MWS was corrected with propensity score based stratification. Continuous and ordinal confounding variables strained the performance of stratified MWS, and this led to unacceptable problems when fitting a large number of strata over a small number of observations. In future stroke research, the stratified MWS is a valid analysis method only when adjustment is needed for a limited number of confounding variables, and when sufficient observations are available to prevent model instability due to empty cells. If it is not possible to keep this ratio of number of strata versus number of observations relatively low, OLR is the superior analysis method. However, propensity score based stratification improves the confounding adjustment performance of MWS, and this should be weighed against the specific limitations of any regression method.

## Data Availability

Code for the computer simulations is provided in the appendices.

